# Neuronal damage and inflammatory biomarkers are associated with the affective and chronic fatigue-like symptoms due to end-stage renal disease

**DOI:** 10.1101/2023.05.03.23289492

**Authors:** Hussein Kadhem Al-Hakeim, Basim Abd Al-Raheem Twaij, Tabarek Hadi Al-Naqeeb, Shatha Rouf Moustafa, Michael Maes

**Affiliations:** Department of Chemistry, Faculty of Science, University of Kufa, Iraq; College of Pharmacy, The Islamic University, Najaf, Iraq; Clinical Analysis Department, College of Pharmacy, Hawler Medical University, Havalan City, Erbil, Iraq; Department of Psychiatry, Faculty of Medicine, Chulalongkorn University, Bangkok, Thailand; Department of Psychiatry, Medical University of Plovdiv, Plovdiv, Bulgaria; Deakin University, IMPACT, the Institute for Mental and Physical Health and Clinical Translation, School of Medicine, Barwon Health, Geelong, Australia; Kyung Hee University, 26 Kyungheedae-ro, Dongdaemun-gu, Seoul 02447, Korea

**Keywords:** neuro-immune, inflammation, mood disorders, biomarkers, neurotoxicity, psychiatry.

## Abstract

**Background:** Many biochemical, immunological, and neuropsychiatric changes are associated with end-stage renal disease (ESRD). Neuronal damage biomarkers such as glial fibrillary acidic protein (GFAP), neurofilament light chain (NFL), S100 calcium-binding protein B (S100B), ionized calcium-binding adaptor molecule-1 (IBA1), and myelin basic protein (MBP) are among the less-studied biomarkers of ESRD.

**Aim:** We examined the associations between these neuro-axis biomarkers, inflammatory biomarkers, e.g., C-reactive protein (CRP), interleukin (IL-6), IL-10, and zinc, copper, and neuropsychiatric symptoms due to ERSD.

**Methods:** ELISA techniques were used to measure serum levels of neuronal damage biomarkers in 70 ESRD patients, and 46 healthy controls.

**Results:** ESRD patients have higher scores of depression, anxiety, fatigue, and physiosomatic symptoms than healthy controls. Aberrations in kidney function tests and the number of dialysis interventions are associated with the severity of depression, anxiety, fibro-fatigue and physiosomatic symptoms, peripheral inflammation, nestin, and NFL. Serum levels of neuronal damage biomarkers (NFL, MBP, and nestin), CRP, and interleukin (IL)-10 are elevated, and serum zinc is decreased in ESRD patients as compared with controls. The neuronal damage biomarkers NFL, nestin, S100B and MBP are associated with the severity of one or more neuropsychiatric symptom domains. Around 50% of the variance in the neuropsychiatric symptoms is explained by NFL, nestin, S00B, copper, and an inflammatory index.

**Conclusions:** The severity of renal dysfunction and/or the number of dialysis interventions may induce peripheral inflammation and, consequently, neurotoxicity to intermediate filament proteins, astrocytes, and the blood-brain barrier, leading to the neuropsychiatric symptoms of ESRD.

## Introduction

End-stage renal disease (ESRD) is accompanied by many biochemical changes, including alterations in immune-inflammatory mediators, such as C-reactive protein (CRP), interleukin (IL-6), and IL-10 (Sinuani et al., 2013, Babaei et al., 2014, Oweis et al., 2021, Su et al., 2017), trace elements, including increased copper and lowered zinc, (Almeida et al., 2020, Dizdar et al., 2020) and oxidative stress biomarkers (Sangeetha Lakshmi et al., 2018, Song et al., 2020). Previous studies have demonstrated that acute ischemic kidney injury (AKI) may cause inflammatory responses in the peripheral blood (Grigoryev et al., 2008). Malfunctions of electrolyte channels and transporters in the injured kidneys may cause abnormalities in sodium, potassium, chloride, and phosphate (Kestenbaum et al., 2005, Einhorn et al., 2009, Barbour et al., 2008), and lowered serum calcium (Timofte et al., 2021).

In association with the biochemical changes, ESRD patients frequently experience physiosomatic symptoms, including chronic fatigue, fibromyalgia, muscular pain, insomnia, headache, cognitive impairments, and affective symptoms, including depression and anxiety (Cohen and Kimmel, 2018, Savitha et al., 2020, Elzeiny and El-Emary, 2023, Ibrahim et al., 2023, Semaan et al., 2018, Qawaqzeh et al., 2023, Khoury et al., 2023, Burdelis and Cruz, 2023, Molfino et al., 2023, Asad et al., 2023). Previous studies showed a high incidence of chronic fatigue (42-89%) and depressive or/and anxiety symptoms (around 50%) in chronic renal disease (Kim et al., 2012, Yoong et al., 2017)(Artom et al., 2014).

There is now evidence that depression, anxiety, chronic fatigue, and physiosomatic symptoms are at least in part mediated through activated immune-inflammatory pathways, including increases in IL-6, IL-10, and CRP (Maes and Carvalho, 2018, Maes et al., 2011b), lowered zinc (Maes et al., 1994, Maes et al., 1997), and increased copper (Ni et al., 2018). Circulating inflammatory mediators and reactive oxygen species may impact brain neuronal circuits, which mediate affective behaviors, thereby causing neuro-affective toxicity (Maes and Carvalho, 2018, Maes et al., 2011b, Morris and Maes, 2013).

Importantly, severe ischemic kidney injury may induce peripheral inflammatory responses that may cause neuro-inflammation and functional abnormalities in neuronal circuits (Liu et al., 2008). Al-Hakeim et al. (2023) observed that, in patients with major depression, indicants of neuronal damage, including elevated levels of plasma neurofilament light chain (NFL), glial fibrillary acidic protein (GFAP), and phosphorylated tau protein 217 (P-tau217), are associated with the “physio-affective phenome” of depression. The latter is a new label to indicate that depressive, anxiety, and physiosomatic symptoms belong to the same latent vector, which therefore is the cause of these symptoms (Maes, 2022b, Al-Hakeim et al., 2023). Moreover, the effects of increased peripheral inflammation and lowered calcium on the physio-affective phenome are mediated by these neuronal damage markers (Al-Hakeim et al., 2023). Importantly, the results of annotation analysis indicate that peripheral mechanisms, including inflammation and lowered calcium, may cause neuronal and astroglial projection toxicity, leading to physio-affective symptoms (Al-Hakeim et al., 2023). Therefore, it could be hypothesized that the immune-inflammatory pathways and lowered calcium levels in ESRD are associated with physio-affective symptoms by causing damage to neuronal cells.

S100 calcium-binding protein B (S100B) and neurofilament light (NFL) are neuronal damage markers (Egea-Guerrero et al., 2013, Kim et al., 2012, Hou et al., 2021) that are associated with ESRD. Serum S100B levels are significantly increased in patients with ESRD and are strongly associated with depressive symptoms and cognitive deficits (Kim et al., 2012, Park et al., 2020). NFL concentrations were marginally increased in ESRD patients, although no significant associations could be established with cognitive functions (Hou et al., 2021). Two other measurable neuronal damage markers that were not examined in ESRD are myelin basic protein (MBP), the main protein of the myelin sheath that plays a key role in maintaining the stability of the myelin sheath (Vyver et al., 2018), and ionized calcium-binding adaptor molecule-1 (IBA1), a microglia/macrophage-specific calcium-binding protein, which is necessary for the phagocytosis of activated microglia (Sasaki et al., 2001, Lafrenaye et al., 2020).

Hence, the current study was conducted to examine whether: a) serum markers of neuronal damage, namely GFAP, NFL, MBP, nestin, IBA1, and S100B are increased in patients with ESRD; b) their levels are independently associated with the severity of the physio-affective phenome of ESRD, comprising depressive, anxiety and physio-somatic symptoms; and c) immune-inflammatory markers such as CRP, IL-6, IL-10, and lower zinc, are associated with the neuronal damage markers.

## Subjects and Methods

### Subjects

The present study involved a sample of seventy patients with ESRD and 46 normal controls. Everyone had experienced an acute kidney injury (AKI) before their current condition, ultimately leading to the development of end-stage renal failure. As a result, all patients were receiving ongoing treatment through the use of continuous dialysis. The study recruited participants from the Dialysis Unit located at Al-Hakeem General Hospital in Najaf Governorate during the period spanning from March to May of 2022. Patient assessment was conducted by considering their complete medical history, including the existence of any systemic disorders. The patients were diagnosed by a senior physician in accordance with the 10^th^ edition of the International Statistical Classification of Diseases and Related Health Problems (ICD-10-CM), specifically under the diagnosis code N18.6. All subjects were under constant medication with folic acid or iron and folate formula (Fefol^®^) along with calcium carbonate, epoetin alfa (Eprex^®^), and heparin. The study’s control group comprised 46 individuals who were in good health and were selected from the same geographical region as the patients. The control group was matched to the patients in terms of age and gender. Neither the patients nor the controls exhibited any lifetime axis-1 diagnoses of prior neuropsychiatric disorders, such as major depressive episodes, anxiety disorders, schizophrenia, bipolar disorder, psycho-organic disorders, substance use disorders (excluding nicotine dependence), and chronic fatigue syndrome. All study participants exhibited no medical conditions apart from ESRD. This includes, but is not limited to, diabetes mellitus, inflammatory diseases, Parkinson’s or Alzheimer’s disease, stroke, oncologic disorders, and (auto)immune disorders.

The patients or their first-degree relatives provided written informed consent. The present study has been approved by the institutional ethics board of the University of Kufa (Document No.2229T / 2022) and the Najaf Health Directorate, Training, and Human Development Center (Document No.20123 / 2022). The research was carried out in compliance with both Iraqi and international regulations on ethics and privacy. Furthermore, it was conducted under the ethical principles outlined in the World Medical Association Declaration of Helsinki. In addition, it is noteworthy that our Institutional Review Board (IRB) adheres to the International Guideline for Human Research Protection, which is mandated by the Declaration of Helsinki, the Belmont Report, the CIOMS Guideline, and the International Conference on Harmonization in Good Clinical Practice (ICH-GCP).

### Clinical assessments

A senior psychiatrist employed a semi-structured interview technique to gather sociodemographic and clinical information. The severity of fibromyalgia and chronic fatigue was assessed by a senior psychiatrist using the FibroFatigue (FF) scale (Zachrisson et al., 2002), while the degree of anxiety was evaluated using the Hamilton Anxiety Rating Scale (HAMA) (Hamilton, 1959). The participants completed the Beck Depression Inventory (BDI)-II rating scale concurrently (Beck et al., 1996). The study employed the total FF, HAMA, and BDI-II rating scales scores and used the items of these scales to derive scores for distinct symptom subdomains. The study measured pure depressive symptoms by computing the total score of BDI-II items feelings of sadness, discouragement about the future, a sense of failure, dissatisfaction, guilt, self-disappointment, self-criticism, suicidal thoughts, crying, loss of interest, difficulty making decisions, a negative self-image, and work inhibition. The construct of pure anxiety was operationalized as the summed score of the HAMA items anxious affect, tension, fears, and anxious conduct exhibited during a structured interview. The present study operationalized pure physiosomatic symptoms as a z-unit-weighted composite score based on FF, HAMA, and BDI-II items: muscle pain, muscle tension, fatigue, autonomic symptoms, gastrointestinal symptoms, headache, and flu-like malaise (all from the FF scale); fatigue and loss of libido (from the BDI-II scale); and somatic muscular, somatic sensory, cardiovascular symptoms, respiratory symptoms, gastrointestinal symptoms, and autonomic symptoms (all from the HAMA scale). The pure FF score was the summation of z scores for muscle pain, muscle tension, fatigue (FF items), somatic muscular (HAMA), and fatigue (BDI).

The diagnosis of Tobacco Use Disorder (TUD) was made following the criteria outlined in the DSM-IV-TR. The calculation of Body Mass Index (BMI) involved the division of body weight in kilograms (kg) by the square of length in meters (m).

### Measurements

Blood samples were collected from the participants after overnight fasting, specifically between 8:00 a.m. and 9:00 a.m. Blood was collected into plain and EDTA tubes. Following the separation process, the serum samples were subsequently allocated into Eppendorf® tubes and kept frozen until thawed for assay. The spectrophotometric measurement of serum albumin, urea, creatinine, total serum protein, copper, and zinc was conducted using kits provided by Spectrum Diagnostics Co. (Cairo, Egypt). The CRP latex slide test manufactured by Spinreact^®^ (Barcelona, Spain) was utilized for conducting CRP assays on human serum. Commercial ELISA sandwich kits provided by Melsin Medical Co. (Jilian, China) and Sunlong Biotech Co., Ltd (Zhejiang, China) were used to measure GFAP, NFL, MBP, nestin, IBA1, S100B, IL-6, and IL-10. We computed two z unit-weighted composite scores: a) an inflammatory composite as z CRP + z IL-6 + z IL-10 – z zinc (z_inflammation); and b) an intermediate filament protein composite as z transformation of nestin (z NES) + z GFAP + z NFL (z_interfilament). The estimated GFR (eGFR) was calculated by using the Modification of Diet in Renal Disease Study equation (Levey et al., 2007): eGFR =175 x (S.Cr)^-1.154^ x (Age)^-0.203^ x 0.742 [if female] x 1.212 [if Black].

### Statistical Analysis

The analysis involved the use of Pearson’s product-moment correlation coefficients to examine the correlations between continuous variables. The study utilized analysis of variance (ANOVA) to investigate the associations among groups and clinical or biomarker data. The evaluation of statistical associations between categorical variables was conducted through contingency table analysis (χ^2^-test). Multiple regression analysis was employed, utilizing a manual method, to evaluate the most important predictors of clinical rating scale scores. Biomarkers were utilized as explanatory variables, while allowing for the potential influence of confounding variables such as age, sex, BMI, and TUD. In addition to utilizing a manual regression technique, an automated technique was also employed, whereby model entry and removal were determined based on p-values of 0.05 and 0.10, respectively. The regression model was evaluated by computing the F-statistic, degrees of freedom, and p-values. Additionally, the proportion of variance accounted for by the model (R^2^), standardized β-coefficients, t-statistics, and p-values were determined for each predictor. Furthermore, an assessment was conducted on the variance inflation factor and tolerance to detect any potential collinearity or multicollinearity issues. The presence of heteroskedasticity was assessed using the White and modified Breusch-Pagan tests for homoscedasticity. All tests were two-tailed, at p=0.05. Principal component (PC) analysis was performed to delineate the first PC extracted from a set of variables (e.g. HAMD. BDI and FF scores). To be accepted as a meaningful and validated construct, all loadings on this first PC should be greater than 0.7, and this PC should account for more than 50% of the total variance in the data. Moreover, the Kaiser-Meyer-Olkin (KMO) metric should be greater than 0.6 (a test for the factorability of the correlation matrix), the anti-image matrix should be adequate, and Bartlett’s test of sphericity should be significant. All statistical analyses were carried out utilizing IBM, SPSS for Windows version 28 (IBM-USA). The primary statistical analyses in this study were the multiple regression analyses delineating the effects of the biomarkers on the physio-affective phenome of ESRD. A priori power analysis (G*Power 3.1.9.4) for linear multiple regression analysis, with an effect size of 0.176 (equivalent to 15% explained variance), alpha=0.05, power=0.8, and 4 covariates, indicates that the minimum sample size should be 73.

## Results

### Construction of PCs of renal functions, dialysis, and clinical domains

Using PCA, we have developed two PCs that reflect kidney function and dialysis characteristics. First, we were able to extract a first PC (PC_kidney) from urea, creatinine, eGFR, TSP, and albumin (KMO=0.878, Bartlett’s test of sphericity χ^2^=368.872, df=10, p=0.001, VE=72.53%, all loadings > 0.783). Second, we extracted the first PC from the number of dialyses per week, the total number of dialyses, and the last day patients underwent dialysis, labeling it PC_dialysis (KMO=0.613, Bartlett’s test of sphericity χ^2^=113.336, df=3, p=0.001, VE=69.79%, all loadings > 0.774). Consequently, we computed a z composite score using PC_kidney + PC_dialysis (labeled: z_diakidney). We were able to extract one validated PC (labeled PC_phenome) from the total FF, HAMA, and BDI scores (KMO=0.730, Bartlett’s test of sphericity χ^2^=311.368, df=3, p=0.001, VE=88.27%, all loadings > 0.908).

### Comparison of demographic and neuropsychiatric scores between ESRD and controls

**Table 1** presents the sociodemographic and clinical data of ESRD patients and the healthy control group. There was no significant difference observed between the two groups in terms of age, sex ratio, BMI, TUD, residency, and employment. The single/married ratio was slightly different between both groups. z_diakidney and all clinical subdomain scores (FF-total, HAM-A total, BDI-total, pure depression, anxiety, physiosomatic, FF, and PC_phenome) were significantly higher in ESRD patients than in controls.

**Table 1.** Demographic and neuropsychiatric rating scale scores of end-stage renal disease (ESRD) patients in comparison with the healthy controls (HC)

| Variable | HC<br>n=46 | ESRD patients<br>n=70 | F / $\chi^2$ | df | p |
| --- | --- | --- | --- | --- | --- |
| Age (years) | 39.90 ± 8.90 | 41.20 ± 12.00 | 0.44 | 1/114 | 0.511 |
| Sex Female/Male | 22/24 | 27/43 | 0.97 | 1 | 0.324 |
| Single/Married | 14/32 | 9/61 | 5.40 | 1 | 0.020 |
| BMI (kg/m<sup>2</sup>) | 24.84 ± 2.54 | 24.06 ± 3.97 | 1.39 | 1/114 | 0.241 |
| Urban / Rural | 40/6 | 55/15 | 1.32 | 1 | 0.251 |
| Employment No/Yes | 27/19 | 40/30 | 0.03 | 1 | 0.868 |
| TUD No/Yes | 40/6 | 65/5 | 1.13 | 1 | 0.289 |
| z_diakidney (z scores) | -1.190 (0.245) | 0.782 (0.249) | MWU |  | <0.001 |
| Total FF score | 6.20 ± 1.80 | 28.10 ± 4.30 | MWU | | <0.001 |
| Total HAMA score | 5.20 ± 1.90 | 21.70 ± 4.40 | MWU | | <0.001 |
| Total BDI-II score | 5.90 ± 1.30 | 21.30 ± 8.50 | MWU | | <0.001 |
| Pure depression score | 4.89 ± 2.48 | 18.66 ± 6.85 | MWU | | <0.001 |
| Pure anxiety score | 1.96 ± 0.99 | 6.86 ± 2.04 | MWU | | <0.001 |
| Pure physiosomatic score (z scores) | -1.16 ± 0.25 | 0.77 ± 0.36 | MWU | | <0.001 |
| Pure FibroFatigue score (z scores) | -1.08 ± 0.37 | 0.71 ± 0.54 | MWU | | <0.001 |
| z_phenome (z score) | -3.024 (0.688) | 1.987 (1.489) | MWU |  | <0.001 |

BMI: Body mass index, TUD: Tobacco use disorder; HAMA: Hamilton Anxiety Rating Scale score; FF: FibroFatigue scale, BDI: Beck's Depression Inventory; z\_diakidney: composite combining kidney function tests and dialysis features; PC\_phenome: the first PC extracted from the total HAMA, BDI-II and FF scores.

### Comparison in serum biomarkers between ESRD and control groups

The biomarker levels in both patient groups and the healthy controls are presented in **Table 2**. Serum levels of NFL, MBP, nestin, z_interfilament, CRP, IL-10, copper, and z_inflammation were significantly higher in patients than in controls. A significant decrease in serum zinc was recorded in ESRD patients compared with controls. No significant differences were established in serum IBA1, S100B, GFAP, or IL-6 between ESRD patients and control groups. After FDR p correction, all significant differences listed in Table 2 remained significant, except MBP (FDR p =0.062).

**Table 2.** Serum biomarkers in end-stage renal disease (ESRD) patients in comparison with healthy controls (HC)

| Variable | HC<br>N=46 | ESRD patients<br>N=70 | F / $\chi^2$ | df | p |
| --- | --- | --- | --- | --- | --- |
| GFAP pg/ml | 132.90±68.40 | 173.80±165.70 | 2.51 | 1/114 | 0.116 |
| NFL ng/ml | 13.30±8.80 | 19.90±12.30 | 1.08 | 1/114 | 0.002 |
| MBP ng/ml | 5.78±2.89 | 7.05±3.50 | 4.14 | 1/114 | 0.044 |
| Nestin pg/ml | 31.50±22.30 | 42.90±28.60 | 5.24 | 1/114 | 0.024 |
| IBA1 pg/ml | 66.00±46.60 | 71.60±52.70 | 0.34 | 1/114 | 0.564 |
| S100B pg/ml | 71.70±37.40 | 82.00±54.00 | 1.28 | 1/114 | 0.260 |
| z_interfilament (z score) | -0.614 ±1.296 | 0.404 ±1.717 | 11.77 | 1/114 | <0.001 |
| CRP mg/l | 0.84±0.53 | 7.51±14.54 | 9.64 | 1/114 | 0.002 |
| IL-6 pg/ml | 5.80±3.60 | 6.20±3.60 | 0.42 | 1/114 | 0.519 |
| IL-10 pg/ml | 6.80±2.50 | 8.10±2.70 | 6.83 | 1/114 | 0.010 |
| Copper mg/l | 0.76±0.28 | 1.16±0.29 | 54.35 | 1/114 | <0.001 |
| Zinc mg/l | 0.81±0.15 | 0.67±0.21 | 16.95 | 1/114 | <0.001 |
| Z_inflammation (z score) | -0.604±0.725 | 0.397±0.960 | 36.29 | 1/114 | <0.001 |

GFAP: Glial fibrillary acidic protein, NFL: Neurofilament light chain, MBP: myelin basic protein, IBA1: ionized calcium-binding adapter molecule 1, CRP: C-reactive protein, IL: interleukin; z\_interfilament: a z composite computed as the sum of the z scores of nestin + NFL + GFAP.

### Correlation matrix

The results of the correlation calculations between clinical scores and the measured biomarkers are presented in **Table 3**. Z-diakidney was significantly correlated with pure depression, anxiety, physiosomatic, and FF scores, z_interfilament, NFL, nestin, and the z_inflammation index. The pure depression score showed a significant correlation with anxiety, physiosomatic and FF scores, z_interfilament, nestin, S100B, and z_inflammation score. The pure anxiety symptom score is correlated significantly with pure physiosomatic and FF scores, z_interfilament, nestin, and z_inflammation. The physiosomatic score showed a significant correlation with the FF score, z_interfilament, NFL, nestin, and z_inflammation. The FF score is significantly correlated with z_interfilament, NFL, and z_inflammation. CRP (r=0.288, p=0.002, n=116) and z_inflammation (r=0.235, p=0.011, n=116) were significantly associated with z_interfilament.

**Table 3.** Correlation matrix between clinical and neuropsychiatric scores with the measured biomarkers

| Parameters | Z_diakidney | Pure depression | Pure anxiety | Pure physiosom | Pure FF |
| --- | --- | --- | --- | --- | --- |
| Z_diakidney | 1 | 0.751** | 0.799** | 0.924** | 0.850** |
| Pure depression | 0.751** | 1 | 0.734** | 0.779** | 0.731** |
| Pure anxiety | 0.799** | 0.734** | 1 | 0.851** | 0.780** |
| Pure physiosomatic | 0.924** | 0.779** | 0.851** | 1 | 0.945** |
| Pure FibroFatigue | 0.850** | 0.731** | 0.780** | 0.945** | 1 |
| z_interfilament | 0.300** | 0.339** | 0.245** | 0.310** | 0.224* |
| GFAP | 0.000 | 0.072 | -0.042 | 0.022 | -0.007 |
| NFL | 0.272** | 0.254** | 0.144 | 0.269** | 0.212* |
| Nestin | 0.218* | 0.229* | 0.298** | 0.217* | 0.162 |
| MBP | 0.182 | 0.236* | 0.120 | 0.163 | 0.073 |
| IBA1 | 0.044 | 0.113 | 0.031 | 0.015 | 0.040 |
| S100B | 0.103 | 0.195* | 0.062 | 0.101 | 0.096 |
| Z_inflammation | 0.467** | 0.573** | 0.501** | 0.459** | 0.410** |

\* $p < 0.05$ ), \*\* $p < 0.01$ ; GFAP: Glial fibrillary acidic protein, z\_diakidney: composite built using z scores of creatinine, urea, estimated glomerular filtration rate, albumin, total serum protein, and number of dialysis sessions, MBP: myelin basic protein, IBA1: Ionized calcium-binding adapter molecule 1, z\_inflammation: z composite computed as the sum of the z transformation of CRP + IL-6 + IL-10 – zinc, z\_interfilament: a z composite computed as the sums of the z scores of nestin + NFL + GFAP

### Prediction of the clinical symptom domains

**Table 4** shows the results of different stepwise multiple regression analyses with clinical symptom scores as dependent variables and biomarkers and confounding variables as independent variables. Firstly (denoted “a” in Table 4), we entered only the single neural variables, and secondly (denoted “b” in Table 4) we also entered z_inflammation, z_interfilament, copper, and background variables. Regression #1.a shows that 14.7% of the variance in the pure depression score was explained by NFL, MBP, and S100B. Regression #1.b shows that the model explained 53.4% of the variance in the pure depression score using z_inflammation, copper, S100B, z_interfilament (all positively associated), education (negatively associated), and sex as explanatory variables. Regression #2.a shows that 8.9% of the variance in the pure anxiety symptoms score was explained by nestin. Regression #2.b shows that a significant part of the pure anxiety variance (39.5%) is explained by z_inflammation, copper, and nestin. Regression #3.a shows that 11.3% of the variance in the pure physiosomatic symptom scores was explained by NFL and nestin. Regression #3.b shows that the model explained 46.0% of the variance in the pure physiosomatic score using z_inflammation, copper, and z_interfilament. **Figure 1** shows the partial regression of the pure physiosomatic symptoms scores on the z_interfilament composite. In Regression #4, 36.7% of the variance in the FF score can be explained by the regression on copper, z_inflammation, and z_interfilament. Regression #5.a examines the effects of the neuronal markers on the phenome and shows that 12.5 % of the variance in PC_phenome was explained by nestin and NFL. Regression #5.b entered neuronal markers as well as other biomarkers and confounding variables and shows that a considerable part of the variance in the phenome score (50.4%) was explained by the regression on z_inflammation, copper, z_interfilament, and S100B. **Figure 2** presents the partial regression of the PC_phenome score on the composite z interfilament score. **Figure 3** shows the partial regression of the phenome score on the z_inflammation composite.

**Figure 1.**
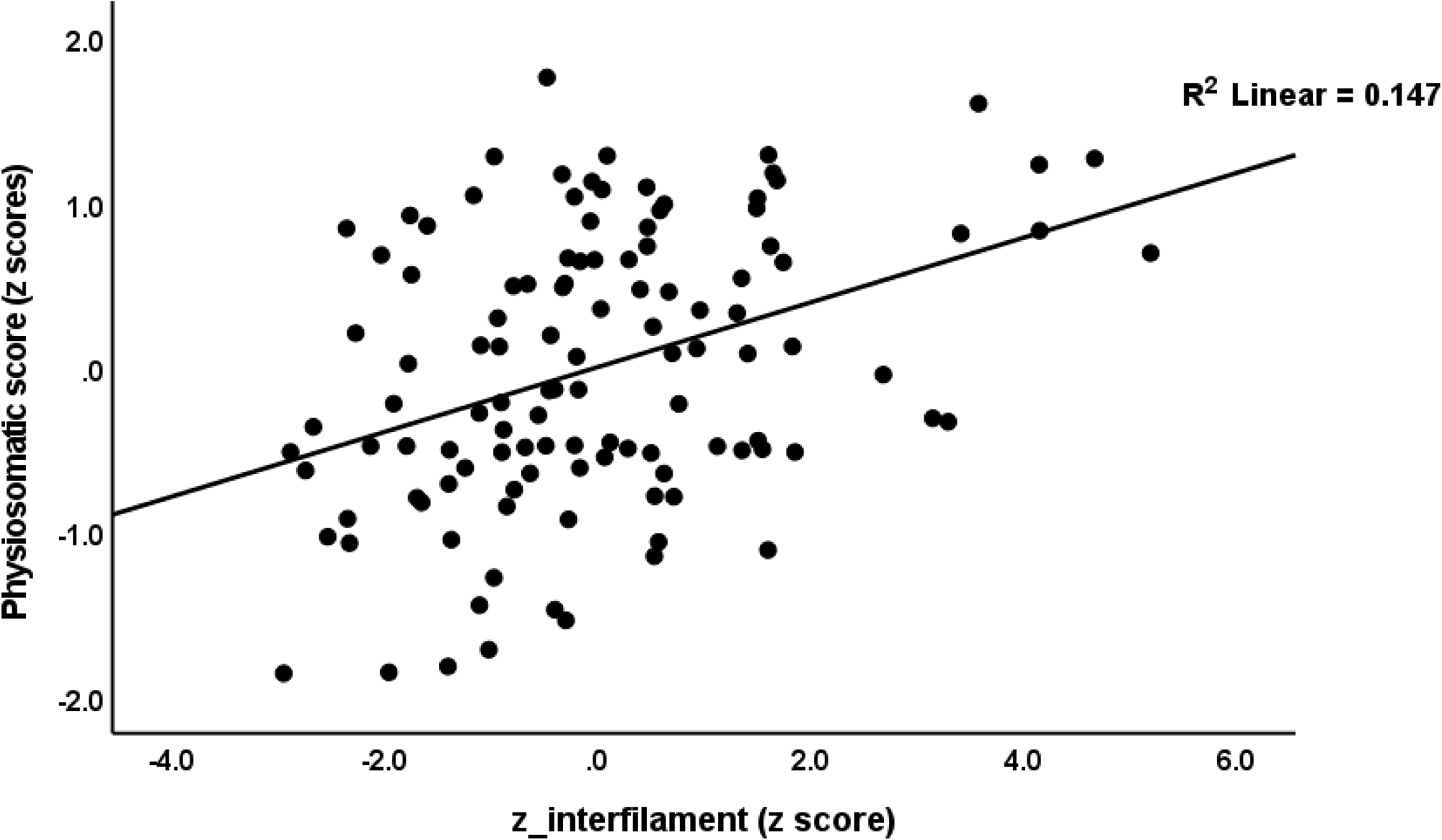
Partial regression of the pure physiosomatic symptoms on z_interfilament, a z composite score based on the interfilament damage biomarkers nestin, glial fibrillary acidic protein, and neurofilament light chain

**Figure 2.**
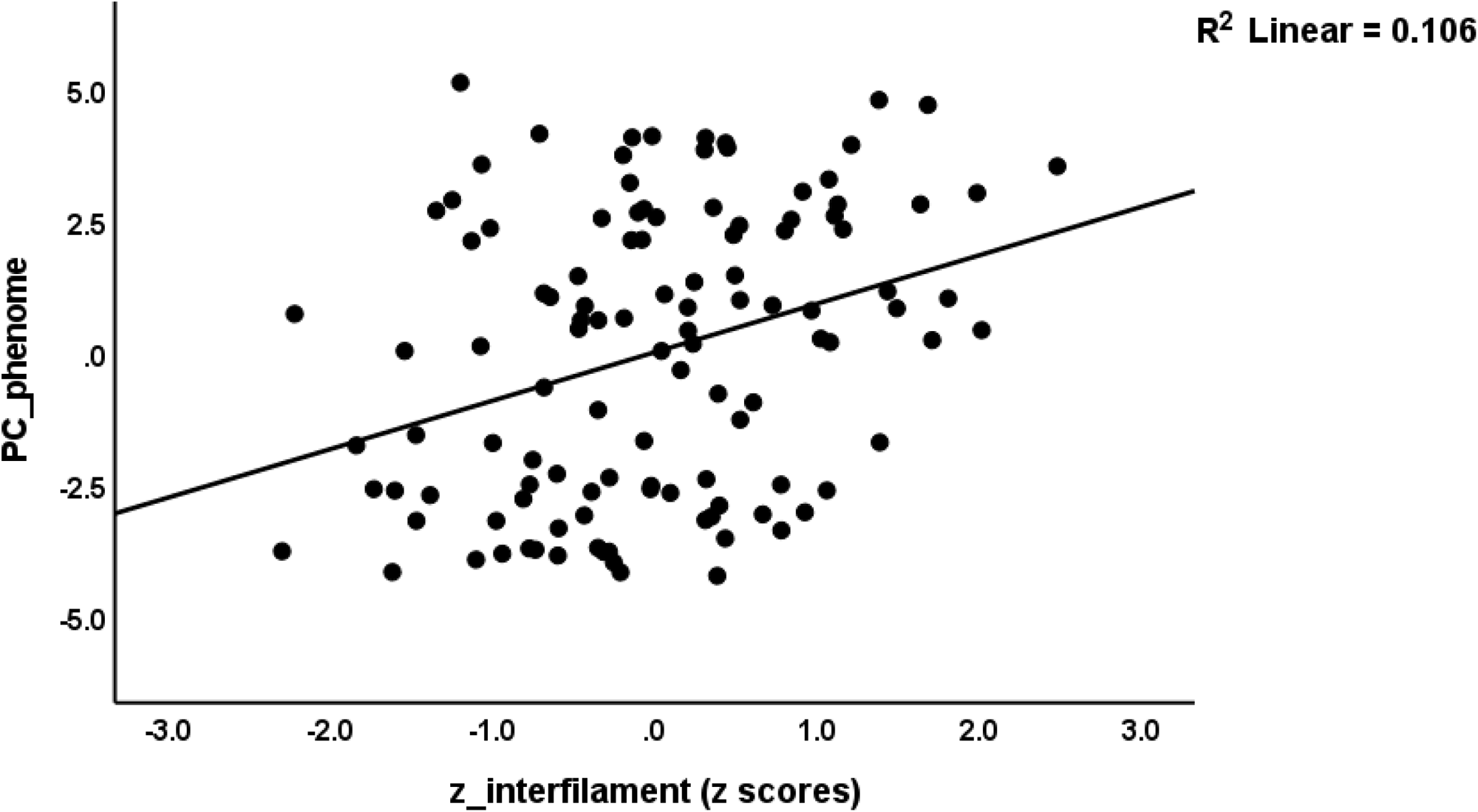
Partial regression of the phenome of end-stage renal disease (comprising depression, anxiety and fibro-fatigue symptoms) on z_interfilament, a z composite score based on the interfilament damage biomarkers nestin, glial fibrillary acidic protein, and neurofilament light chain

**Figure 3.**
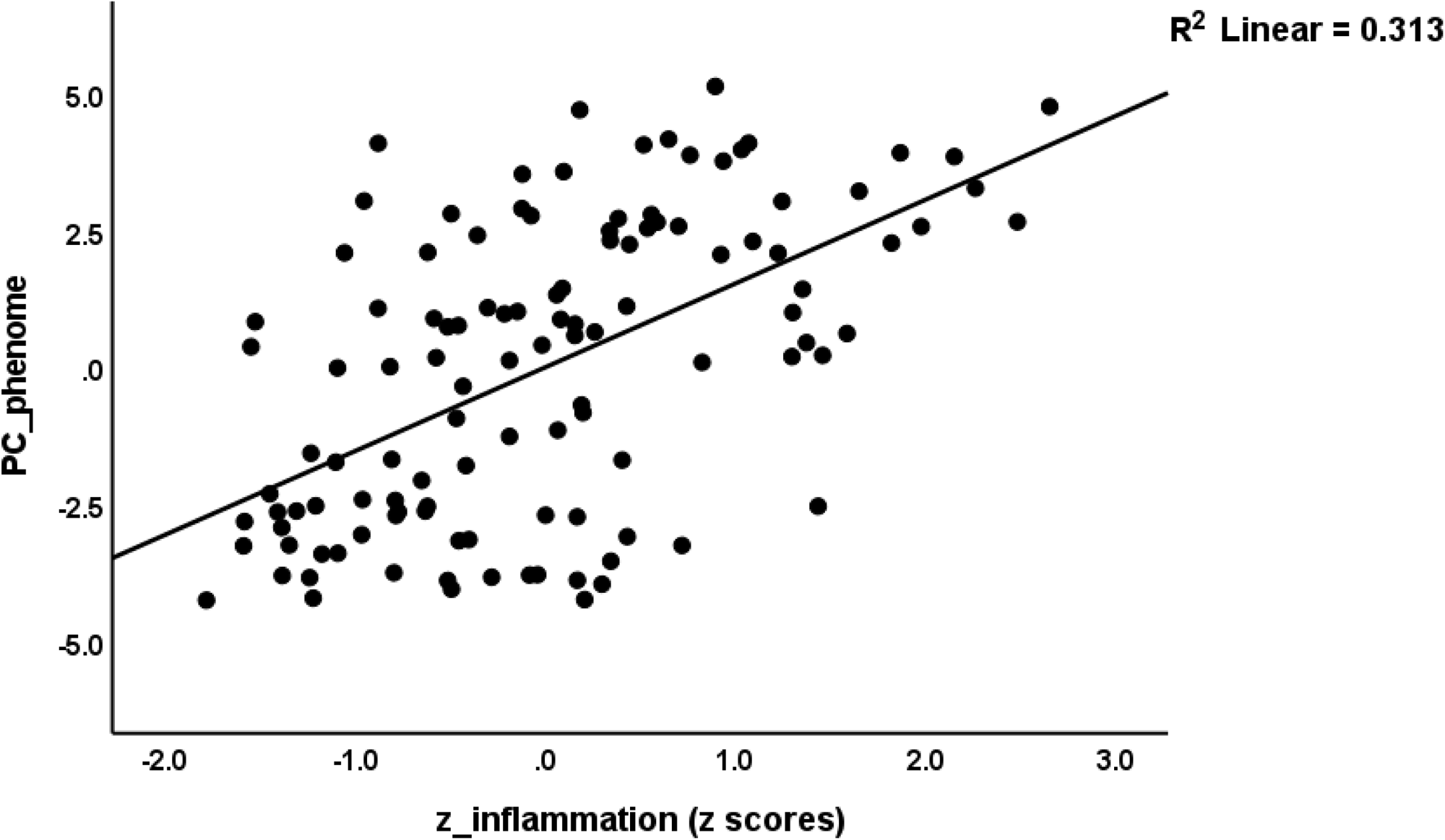
Partial regression of the of the phenome of end-stage renal disease (comprising depression, anxiety and fibro-fatigue symptoms) on z_inflammation, a composite based on 4 inflammatory biomarkers.

**Table 4.**
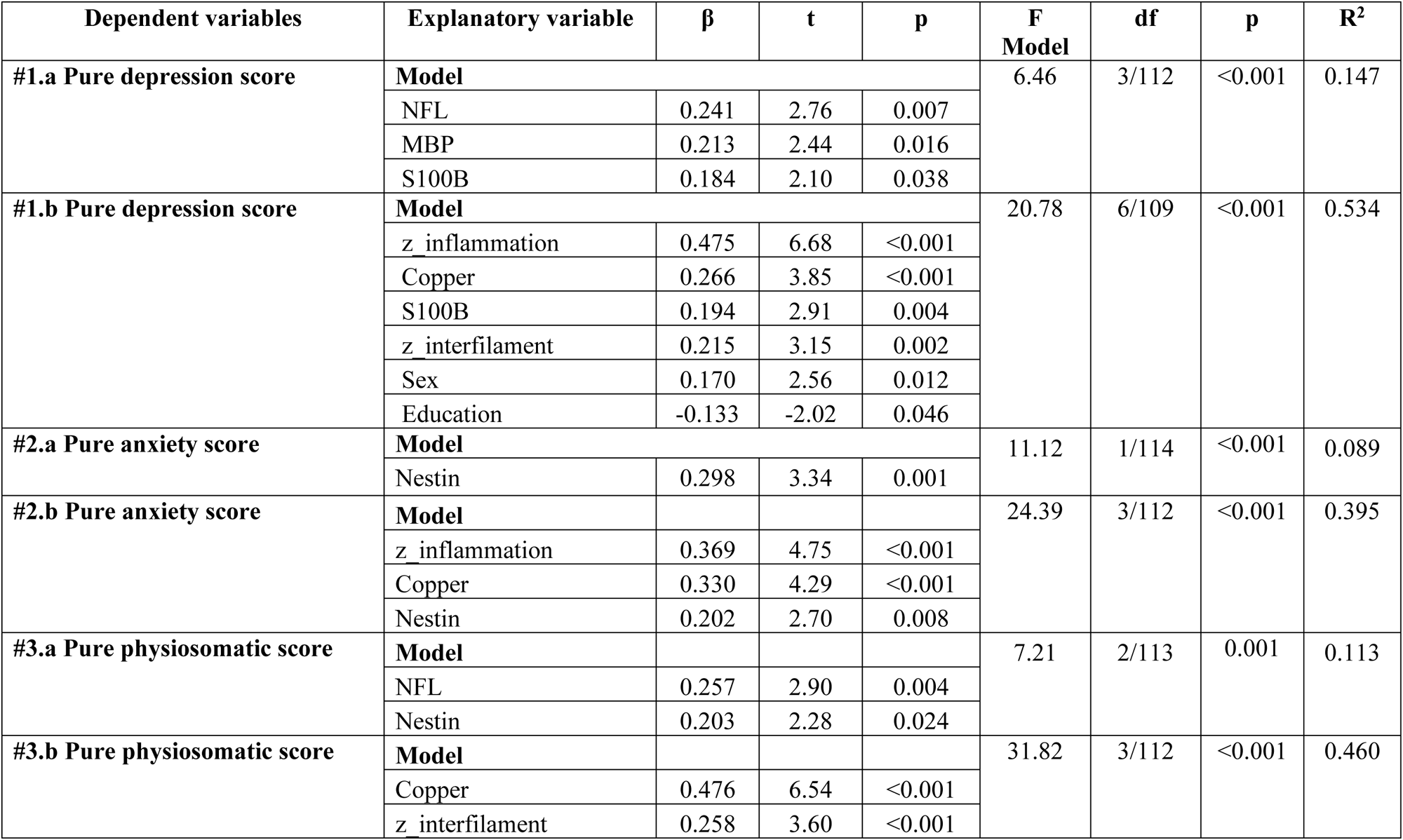
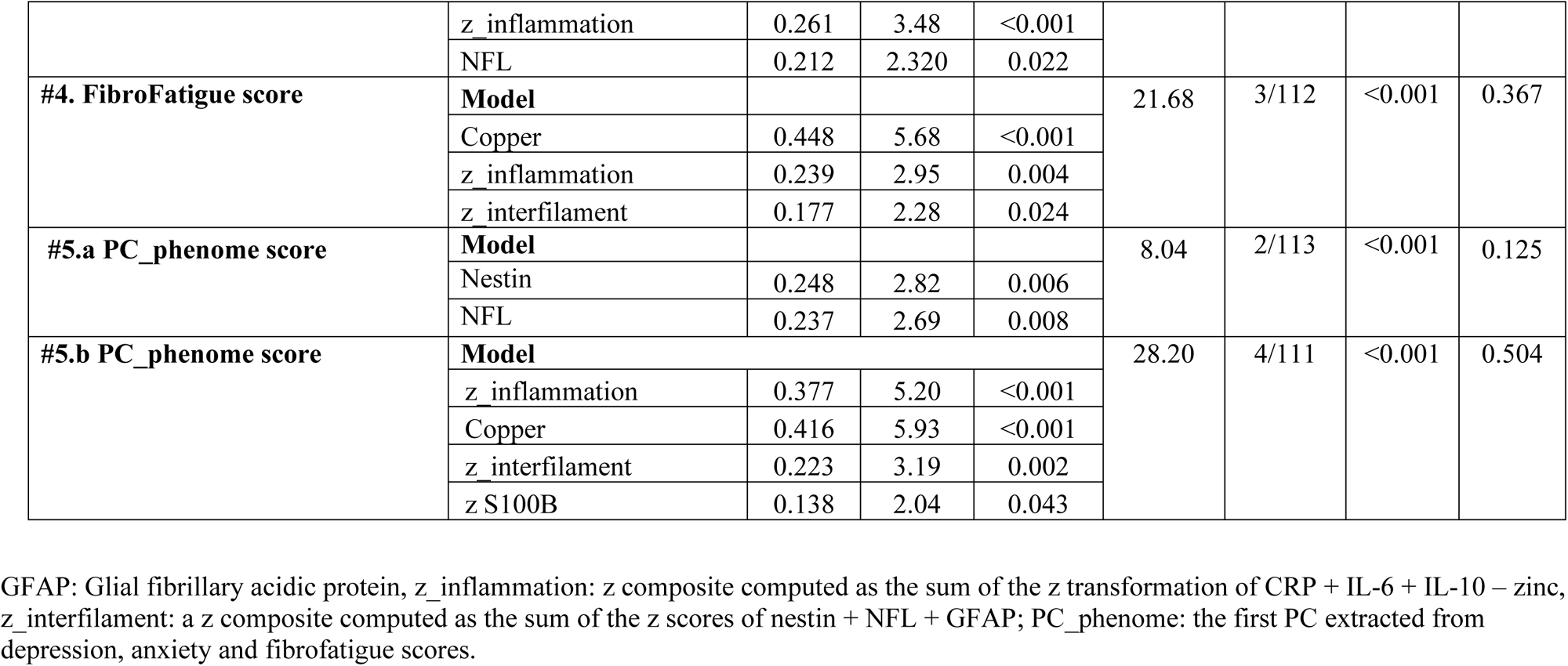
Results of multiple regression analyses with clinical subdomain scores of end-stage renal disease as dependent variables

| **Dependent variables** | **Explanatory variable** | **$\beta$** | **t** | **p** | **F Model** | **df** | **p** | **R<sup>2</sup>** |
| --- | --- | --- | --- | --- | --- | --- | --- | --- |
| **#1.a Pure depression score** | **Model** |  |  |  | 6.46 | 3/112 | <0.001 | 0.147 |
|  | NFL | 0.241 | 2.76 | 0.007 |  |  |  |  |
|  | MBP | 0.213 | 2.44 | 0.016 |  |  |  |  |
|  | S100B | 0.184 | 2.10 | 0.038 |  |  |  |  |
| **#1.b Pure depression score** | **Model** |  |  |  | 20.78 | 6/109 | <0.001 | 0.534 |
|  | z_inflammation | 0.475 | 6.68 | <0.001 |  |  |  |  |
|  | Copper | 0.266 | 3.85 | <0.001 |  |  |  |  |
|  | S100B | 0.194 | 2.91 | 0.004 |  |  |  |  |
|  | z_interfilament | 0.215 | 3.15 | 0.002 |  |  |  |  |
|  | Sex | 0.170 | 2.56 | 0.012 |  |  |  |  |
|  | Education | -0.133 | -2.02 | 0.046 |  |  |  |  |
| **#2.a Pure anxiety score** | **Model** |  |  |  | 11.12 | 1/114 | <0.001 | 0.089 |
|  | Nestin | 0.298 | 3.34 | 0.001 |  |  |  |  |
| **#2.b Pure anxiety score** | **Model** |  |  |  | 24.39 | 3/112 | <0.001 | 0.395 |
|  | z_inflammation | 0.369 | 4.75 | <0.001 |  |  |  |  |
|  | Copper | 0.330 | 4.29 | <0.001 |  |  |  |  |
|  | Nestin | 0.202 | 2.70 | 0.008 |  |  |  |  |
| **#3.a Pure physiosomatic score** | **Model** |  |  |  | 7.21 | 2/113 | 0.001 | 0.113 |
|  | NFL | 0.257 | 2.90 | 0.004 |  |  |  |  |
|  | Nestin | 0.203 | 2.28 | 0.024 |  |  |  |  |
| **#3.b Pure physiosomatic score** | **Model** |  |  |  | 31.82 | 3/112 | <0.001 | 0.460 |
|  | Copper | 0.476 | 6.54 | <0.001 |  |  |  |  |
|  | z_interfilament | 0.258 | 3.60 | <0.001 |  |  |  |  |
|  | z_inflammation | 0.261 | 3.48 | <0.001 |  |  |  |  |
|  | NFL | 0.212 | 2.320 | 0.022 |  |  |  |  |
| **#4. FibroFatigue score** | **Model** |  |  |  | 21.68 | 3/112 | <0.001 | 0.367 |
|  | Copper | 0.448 | 5.68 | <0.001 |  |  |  |  |
|  | z_inflammation | 0.239 | 2.95 | 0.004 |  |  |  |  |
|  | z_interfilament | 0.177 | 2.28 | 0.024 |  |  |  |  |
| **#5.a PC_phenome score** | **Model** |  |  |  | 8.04 | 2/113 | <0.001 | 0.125 |
|  | Nestin | 0.248 | 2.82 | 0.006 |  |  |  |  |
|  | NFL | 0.237 | 2.69 | 0.008 |  |  |  |  |
| **#5.b PC_phenome score** | **Model** |  |  |  | 28.20 | 4/111 | <0.001 | 0.504 |
|  | z_inflammation | 0.377 | 5.20 | <0.001 |  |  |  |  |
|  | Copper | 0.416 | 5.93 | <0.001 |  |  |  |  |
|  | z_interfilament | 0.223 | 3.19 | 0.002 |  |  |  |  |
|  | z S100B | 0.138 | 2.04 | 0.043 |  |  |  |  |

GFAP: Glial fibrillary acidic protein, z\_inflammation: z composite computed as the sum of the z transformation of CRP + IL-6 + IL-10 – zinc, z\_interfilament: a z composite computed as the sum of the z scores of nestin + NFL + GFAP; PC\_phenome: the first PC extracted from depression, anxiety and fibrofatigue scores.

## Discussion

### The physio-affective phenome of ESRD

The first major finding of the current study is the highly significant increase in the total and subdomain scores of neuropsychiatric symptoms (anxiety, depression, and FF) in ESRD, and that we were able to extract one principal component from those symptom domain scores. These results extend those of previous publications showing a high prevalence of neuropsychiatric disorders among ESRD patients including depression (Elzeiny and El-Emary, 2023, Ibrahim et al., 2023, Cohen and Kimmel, 2018, Savitha et al., 2020), anxiety (Semaan et al., 2018, Qawaqzeh et al., 2023, Khoury et al., 2023), and fatigue (Burdelis and Cruz, 2023, Molfino et al., 2023). In those patients, fatigue (Bossola et al., 2015), depression (Lopes et al., 2002), and anxiety (Schouten et al., 2019) are considered to be independent risk factors for increased number of hospitalizations, morbidity, and mortality. Affective disorders due to ESRD are well-known risk factors that impact disabilities, the health, and the quality of life of patients on hemodialysis treatment (Cukor et al., 2007). These studies show that to lower morbidity and mortality in ESRD patients, neuropsychiatric symptoms need to be evaluated, integrated into the severity assessment, and considered while treating ESRD patients.

In addition, our findings indicate that deteriorating kidney function and the number of dialysis treatments are substantially associated with the severity of the physio-affective phenome. Therefore, it is possible to conclude that neurotoxic factors related to ESRD and/or dialysis treatment may be responsible for the interrelated increases in depression, anxiety, and fatigue-physiosomatic symptoms.

### Biomarkers of ESRD

The second major finding of our study is that NFL, nestin, a composite based on the three intermediate filament proteins (NFL, nestin, and GFAP), CRP, IL-10, and copper are increased, and zinc decreased in patients with ESRD as compared with the control group. In addition, deterioration of kidney function and an increase in the number of dialysis interventions were significantly associated with the intermediate filament composite, particularly NFL and nestin.

Increased NFL, a neuro-axonal damage biomarker (Watanabe et al., 2019), reflects ongoing and acute disease processes in patients with neuroinflammatory illnesses (Cantó et al., 2019, Srpova et al., 2021). As such, our results indicate a potential connection between increasing neurotoxicity due to renal failure, aberrations in neuronal cell communication and cell structure, and astrocyte functions as indicated by increased NFL levels (Wang et al., 2017b, Maes, 2022a). Nestin is another intermediate filament protein that is expressed by neuronal progenitor cells, including those in the adult brain; and that mediates radial growth of the axon (Hendrickson et al., 2011). Nestin-expressing neurons are mainly observed in brain regions involved in cognitive functions suggesting that nestin plays a key role in neuroplasticity. Re-expression of nestin in reactive astrocytes and glial scars may be triggered by injuries such as traumatic and ischemic brain lesions (Kernie et al., 2001, Liberto et al., 2004, Gilyarov and physiology, 2008). However, nestin is not exclusively expressed in neurons but is also expressed by kidney cells (Su et al., 2008), especially in the hypoxic regions of the kidney and following tubulointerstitial injuries (Sakairi et al., 2007, Tomioka et al., 2010). Increased expression of nestin has been reported during the regeneration of injuries to skeletal muscles (Oikawa et al., 2010).

Another intermediate filament found in the cytoskeleton of astrocytes is GFAP, which regulates astrocyte activity, cell structure, and communication between cells (Nedergaard et al., 2003, Wang et al., 2017a, Maes et al., 2023). In response to CNS damage and reactive astrogliosis, GFAP expression is elevated. According to Halford et al. (2017), damaged astrocytes can leak GFAP into the neuronal regions (Halford et al., 2017), and GFAP may be translocated into the peripheral circulation through the breakdown of the BBB (Hol and Pekny, 2015, Yang and Wang, 2015). So, the presence of GFAP in plasma may indicate increased neurotoxicity, neuronal, and glial cell damage and may even predict astrocyte cell death (Yang and Wang, 2015, Halford et al., 2017). Although increases in the two other intermediate filament proteins were observed in ESRD, no significant changes could be established in serum GFAP.

The increase in MBP in ESRD patients has not previously been studied. In cases of traumatic brain injury, benign and malignant intracranial tumors, CNS infections, and cerebrovascular accidents, serum levels of MBP are typically elevated. Increased concentrations serve as a specific indicator of demyelination (Wąsik et al., 2020). MBP is extremely basic and carries highly positive charges (Harauz et al., 2009) and it directly interacts with acidic components on the external side of the neuronal membrane to generate neurotoxicity. Therefore, it is probable that MBP plays a role in the etiology of CNS illnesses characterized by myelin destruction (Zhang et al., 2014).

### Increased neuro-axis biomarkers are associated with the physio-affective phenome

The third major finding of this study is that the intermediate filament-based composite, and NFL, MBP, and S100B are significantly associated with the different symptom domains (depression, anxiety, physiosomatic, and FF scores) as well as with the physio-affective phenome of ESRD. In addition, the serum levels of intermediate filaments are elevated in ESRD and, in conjunction with S100B, are associated with the severity of neuropsychiatric symptoms, independent of the effects of inflammatory mediators and copper.

S100B is predominantly expressed by astrocytes and plays a role in neurite extension, astrocytosis, and axonal proliferation; however, its expression is increased in the adult brain following neuronal damage (Gerlach et al., 2006). During brain injury, astrocytes secrete S100B that may leak into the systemic circulation, which explains the higher serum levels in patients with neurodegenerative disorders, brain injuries, and breakdown of the blood-brain barrier (BBB) (Wang and Bordey, 2008, Chaves et al., 2010, Krishnan et al., 2020, Langeh and Singh, 2021).

Our results extend those of previous reports. For example, increased serum levels of neuronal damage biomarkers, namely PDGFR, NFL, GFAP, and phosphorylated tau protein 217, were observed in MDD patients as compared with controls (Al-Hakeim et al., 2023). Serum levels of NFL and S100B are significantly higher in MDD than in controls (Arora et al., 2019, Chen et al., 2022). NFL serum levels are inversely related to cognitive functions in depressed patients (Bavato et al., 2021). Furthermore, serum GFAP levels may rise with increasing MDD severity (Steinacker et al., 2021), and elevations in plasma GFAP were observed in individuals with neuropsychiatric illnesses who lack brain radiological abnormalities (Esnafoglu et al., 2017).

Interestingly, we detected that the inflammatory composite, indicating an acute phase response in our ESRD patients (e.g., increased CRP and lowered zinc) was significantly associated with increases in the intermediate filament proteins. This is in agreement with a previous report that increased tryptophan catabolites (TRYCATs) in patients undergoing dialysis are associated with depression and fatigue (Malhotra et al., 2017). TRYCATs are induced by pro-inflammatory cytokines and many TRYCATs have neurotoxic effects (Almulla et al., 2022). Previous work also found that severe AKI induces inflammation in association with functional changes in the brain (Liu et al., 2008). Previously, we observed that, in major depression, inflammatory mediators were significantly associated with an increase in GFAP, NFL, and phosphorylated tau protein 217 (Al-Hakeim et al., 2023). The associations between intermediate filament proteins and S100B, inflammatory mediators, and neuropsychiatric symptoms due to ESRD suggest that the severity of kidney dysfunctions and/or the number of dialysis interventions cause inflammation and consequently neuro-affective toxicity leading to neuropsychiatric symptoms.

## Limitations

This study would have been more interesting if we had also measured nitro-oxidative damage markers, including those of lipid peroxidation and hypernitrosylation (Maes et al., 2011a). Our study has a case-control design, and therefore no firm conclusions can be drawn on causality.

## Conclusion

Serum levels of intermediate filament proteins such as NFL, nestin, and S100B are correlated with the severity of neuropsychiatric symptoms associated with ESRD. Overall, the findings suggest that the severity of kidney dysfunction and/or the number of dialysis interventions induce inflammation and, as a result, neuro-affective toxicity, which results in neuropsychiatric symptoms.

## Data Availability

The corresponding author (MM) will make the dataset available upon reasonable request and after the authors have fully exploited the dataset generated and/or analyzed during the current study.

## Acknowledgment

The authors express gratitude to the personnel of the Dialysis Unit at Al-Hakeem General Hospital for their assistance in sample collection, as well as to Asia Clinical Laboratory in Najaf for conducting hematological and biochemical analyses.

## Author contributions

All authors involved in the preparation of the manuscript have made contributions.

## References

Al-Hakeim, H. K., Al-Naqeeb, T. H., Almulla, A. F. & Maes, M. J. J. O. A. D. 2023. The physio-affective phenome of major depression is strongly associated with biomarkers of astroglial and neuronal projection toxicity which in turn are associated with peripheral inflammation, insulin resistance and lowered calcium. 331, 300–312.

Almeida, A., Gajewska, K., Duro, M., Costa, F. & Pinto, E. 2020. Trace element imbalances in patients undergoing chronic hemodialysis therapy - Report of an observational study in a cohort of Portuguese patients. J Trace Elem Med Biol, 62, 126580.

Almulla, A. F., Supasitthumrong, T., Amrapala, A., Tunvirachaisakul, C., Jaleel, A.-K. K. A., Oxenkrug, G., Al-Hakeim, H. K. & Maes, M. J. J. O. A. S. D. 2022. The tryptophan catabolite or kynurenine pathway in Alzheimer’s disease: a systematic review and meta-analysis. 1–15.

Arora, P., Sagar, R., Mehta, M., Pallavi, P., Sharma, S. & Mukhopadhyay, A. K. J. I. J. O. P. 2019. Serum S100B levels in patients with depression. 61, 70.

Artom, M., Moss-Morris, R., Caskey, F. & Chilcot, J. 2014. Fatigue in advanced kidney disease. Kidney Int, 86, 497–505.

Asad, H. N., Al-Hakeim, H. K., Moustafa, S. R. & Maes, M. 2023. A Causal-Pathway Phenotype of Chronic Fatigue Syndrome due to Hemodialysis in Patients with End-Stage Renal Disease. CNS Neurol Disord Drug Targets, 22, 191–206.

Babaei, M., Dashti, N., Lamei, N., Abdi, K., Nazari, F., Abbasian, S. & Gerayeshnejad, S. 2014. Evaluation of plasma concentrations of homocysteine, IL-6, TNF-alpha, hs-CRP, and total antioxidant capacity in patients with end-stage renal failure. Acta Med Iran, 52, 893–8.

Barbour, S. J., Er, L., Djurdjev, O., Karim, M. A. & Levin, A. 2008. The prevalence of hematologic and metabolic abnormalities during chronic kidney disease stages in different ethnic groups. Kidney Int, 74, 108–14.

Bavato, F., Cathomas, F., Klaus, F., Gutter, K., Barro, C., Maceski, A., Seifritz, E., Kuhle, J., Kaiser, S. & Quednow, B. B. 2021. Altered neuroaxonal integrity in schizophrenia and major depressive disorder assessed with neurofilament light chain in serum. J Psychiat Res, 140, 141–148.

Beck, A. T., Steer, R. A. & Brown, G. 1996. Beck depression inventory–II. Psychol Assess.

Bossola, M., Di stasio, E., Antocicco, M., Panico, L., Pepe, G. & Tazza, L. 2015. Fatigue is associated with increased risk of mortality in patients on chronic hemodialysis. Nephron, 130, 113–118.

Burdelis, R. E. M. & Cruz, F. J. S. M. 2023. Prevalence and predisposing factors for fatigue in patients with chronic renal disease undergoing hemodialysis: a cross-sectional study. Sao Paulo Med J, 141, e2022127.

Cantó, E., Barro, C., Zhao, C., Caillier, S. J., Michalak, Z., Bove, R., Tomic, D., Santaniello, A., Häring, D. A. & Hollenbach, J. 2019. Association between serum neurofilament light chain levels and long-term disease course among patients with multiple sclerosis followed up for 12 years. JAMA neurology, 76, 1359–1366.

Chaves, M. L., Camozzato, A. L., Ferreira, E. D., Piazenski, I., Kochhann, R., Dall’igna, O., Mazzini, G. S., Souza, D. O. & Portela, L. V. J. J. O. N. 2010. Serum levels of S100B and NSE proteins in Alzheimer’s disease patients. 7, 1–7.

Chen, M.-H., Liu, Y.-L., Kuo, H.-W., Tsai, S.-J., Hsu, J.-W., Huang, K.-L., Tu, P.-C. & Bai, Y.-M. 2022. Neurofilament light chain is a novel biomarker for major depression and related executive dysfunction. Int J Neuropsychopharmacol, 25, 99–105.

Cohen, S. D. & Kimmel, P. L. J. C. J. O. T. A. S. O. N. 2018. Management of Nonadherence in ESKD Patients. CJN. 13331117.

Cukor, D., Cohen, S. D., Peterson, R. A. & Kimmel, P. L. J. J. O. T. A. S. O. N. 2007. Psychosocial aspects of chronic disease: ESRD as a paradigmatic illness. J Am Soc Nephrol, 18, 3042–3055.

Dizdar, O. S., Yıldız, A., Gul, C. B., Gunal, A. I., Ersoy, A. & Gundogan, K. 2020. The effect of hemodialysis, peritoneal dialysis and renal transplantation on nutritional status and serum micronutrient levels in patients with end-stage renal disease; Multicenter, 6-month period, longitudinal study. J Trace Elem Med Biol, 60, 126498.

Egea-Guerrero, J. J., Murillo-Cabezas, F., Gordillo-Escobar, E., Rodríguez-Rodríguez, A., Enamorado-Enamorado, J., Revuelto-Rey, J., Pacheco-Sánchez, M., León-Justel, A., Domínguez-Roldán, J. M. & Vilches-Arenas, A. J. J. O. N. 2013. S100B protein may detect brain death development after severe traumatic brain injury. J Neurotrauma, 30, 1762–1769.

Einhorn, L. M., Zhan, M., Hsu, V. D., Walker, L. D., Moen, M. F., Seliger, S. L., Weir, M. R. & Fink, J. C. 2009. The frequency of hyperkalemia and its significance in chronic kidney disease. Arch Intern Med, 169, 1156–62.

Elzeiny, H. H. & El-Emary, F. M. 2023. Efficacy of Psychological Interventions in Reducing the Prevalence and Intensity of Depression in Patients with Long-Term Hemodialysis. Egyp J Health Care, 14, 132–143.

Esnafoglu, E., Ayyıldız, S. N., Cırrık, S., Erturk, E. Y., Erdil, A., Daglı, A. & Noyan, T. 2017. Evaluation of serum Neuron-specific enolase, S100B, myelin basic protein and glial fibrilliary acidic protein as brain specific proteins in children with autism spectrum disorder. Int J Dev Neurosci, 61, 86–91.

Gerlach, R., Demel, G., König, H.-G., Gross, U., Prehn, J., Raabe, A., Seifert, V. & Kögel, D. J. N. 2006. Active secretion of S100B from astrocytes during metabolic stress. 141, 1697–1701.

Gilyarov, A. J. N. & Physiology, B. 2008. Nestin in central nervous system cells. Neurosci Behav Physiol, 38, 165–169.

Grigoryev, D. N., Liu, M., Hassoun, H. T., Cheadle, C., Barnes, K. C. & Rabb, H. 2008. The local and systemic inflammatory transcriptome after acute kidney injury. J Am Soc Nephrol, 19, 547–558.

Halford, J., Shen, S., Itamura, K., Levine, J., Chong, A. C., Czerwieniec, G., Glenn, T. C., Hovda, D. A., Vespa, P., Bullock, R. J. J. O. C. B. F. & METABOLISM 2017. New astroglial injury-defined biomarkers for neurotrauma assessment. 37, 3278–3299.

Hamilton, M. 1959. The assessment of anxiety states by rating. Br J Med Psychol, 32, 50–5.

Harauz, G., Ladizhansky, V. & Boggs, J. M. 2009. Structural polymorphism and multifunctionality of myelin basic protein. Biochemistry, 48, 8094–8104.

Hendrickson, M. L., Rao, A. J., Demerdash, O. N. & Kalil, R. E. 2011. Expression of nestin by neural cells in the adult rat and human brain. PLoS One, 6, e18535.

Hol, E. M. & Pekny, M. J. C. O. I. C. B. 2015. Glial fibrillary acidic protein (GFAP) and the astrocyte intermediate filament system in diseases of the central nervous system. 32, 121–130.

Hou, Y. C., Huang, C. L., Lu, C. L., Zheng, C. M., Lin, Y. F., Lu, K. C., Chung, Y. L. & Chen, R. M. 2021. The Role of Plasma Neurofilament Light Protein for Assessing Cognitive Impairment in Patients With End-Stage Renal Disease. Front Aging Neurosci, 13, 657794.

Ibrahim, M. E.-T., Ali el-sayed, S. S. & Abd-Elmaksoud, S. F. J. B. J. O. A. S. 2023. Depression among Egyptian End Stage Renal Disease Patients Under Maintenance Hemodialysis. Benha J Appl Sci.

Kernie, S. G., Erwin, T. M. & Parada, L. F. J. J. O. N. R. 2001. Brain remodeling due to neuronal and astrocytic proliferation after controlled cortical injury in mice. J Neurosci Res, 66, 317–326.

Kestenbaum, B., Sampson, J. N., Rudser, K. D., Patterson, D. J., Seliger, S. L., Young, B., Sherrard, D. J. & Andress, D. L. 2005. Serum phosphate levels and mortality risk among people with chronic kidney disease. J Am Soc Nephrol, 16, 520–8.

Khoury, R., Ghantous, Z., Ibrahim, R., Ghossoub, E., Madaghjian, P., Karam, E., Karam, G., Fares, N. & Karam, S. 2023. Anxiety, depression and post-traumatic stress disorder in patients on hemodialysis in the setting of the pandemic, inflation, and the Beirut blast: a cross-sectional study. BMC Psychiatry, 23, 1–10.

Kim, J.-K., Kim, S. G., Kim, H. J. & Song, Y. R. 2012. Serum S100B protein is associated with depressive symptoms in patients with end-stage renal disease. Clinical Biochemistry, 45, 1573–1577.

Krishnan, A., Wu, H. & Venkataraman, V. J. G. H. D. 2020. Astrocytic S100B, blood-brain barrier and neurodegenerative diseases.

Lafrenaye, A. D., Mondello, S., Wang, K. K., Yang, Z., Povlishock, J. T., Gorse, K., Walker, S., Hayes, R. L. & Kochanek, P. M. J. S. R. 2020. Circulating GFAP and Iba-1 levels are associated with pathophysiological sequelae in the thalamus in a pig model of mild TBI. Sci Rep, 10, 1–17.

Langeh, U. & Singh, S. J. C. N. 2021. Targeting S100B protein as a surrogate biomarker and its role in various neurological disorders. 19, 265–277.

Levey, A. S., Coresh, J., Greene, T., Marsh, J., Stevens, L. A., Kusek, J. W., Van lente, F. & COLLABORATION, C. K. D. E. 2007. Expressing the Modification of Diet in Renal Disease Study equation for estimating glomerular filtration rate with standardized serum creatinine values. Clin Chem, 53, 766–772.

Liberto, C. M., Albrecht, P., Herx, L., Yong, V. & Levison, S. J. J. O. N. 2004. Pro-regenerative properties of cytokine-activated astrocytes. J Neurochem, 89, 1092–1100.

Liu, M., Liang, Y., Chigurupati, S., Lathia, J. D., Pletnikov, M., Sun, Z., Crow, M., Ross, C. A., Mattson, M. P. & Rabb, H. 2008. Acute kidney injury leads to inflammation and functional changes in the brain. J Am Soc Nephrol, 19, 1360–70.

Lopes, A. A., Bragg, J., Young, E., Goodkin, D., Mapes, D., Combe, C., Piera, L., Held, P., Gillespie, B. & Port, F. K. J. K. I. 2002. Depression as a predictor of mortality and hospitalization among hemodialysis patients in the United States and Europe. Kidney int, 62, 199–207.

Maes, M. 2022a. Precision Nomothetic Medicine in Depression Research: A New Depression Model, and New Endophenotype Classes and Pathway Phenotypes, and A Digital Self. J Pers Med, 12.

Maes, M. & Carvalho, A. F. J. M. N. 2018. The compensatory immune-regulatory reflex system (CIRS) in depression and bipolar disorder. 55, 8885–8903.

Maes, M., D’haese, P., Scharpe, S., D’hondt, P., Cosyns, P. & De broe, M. J. J. O. A. D. 1994. Hypozincemia in depression. 31, 135–140.

Maes, M., Galecki, P., Chang, Y. S., Berk, M. J. P. I. N.-P. & Psychiatry, B. 2011a. A review on the oxidative and nitrosative stress (O&NS) pathways in major depression and their possible contribution to the (neuro) degenerative processes in that illness. 35, 676–692.

Maes, M., Ruckoanich, P., Chang, Y. S., Mahanonda, N., Berk, M. J. P. I. N.-P. & Psychiatry, B. 2011b. Multiple aberrations in shared inflammatory and oxidative & nitrosative stress (IO&NS) pathways explain the co-association of depression and cardiovascular disorder (CVD), and the increased risk for CVD and due mortality in depressed patients. 35, 769–783.

Maes, M., Thisayakorn, P., Thipakorn, Y., Tantavisut, S., Sirivichayakul, S. & Vojdani, A. 2023. Reactivity to neural tissue epitopes, aquaporin 4 and heat shock protein 60 is associated with activated immune-inflammatory pathways and the onset of delirium following hip fracture surgery. Eur Geriatr Med, 14, 99–112.

Maes, M., Vandoolaeghe, E., Neels, H., Demedts, P., Wauters, A., Meltzer, H. Y., Altamura, C. & Desnyder, R. J. B. P. 1997. Lower serum zinc in major depression is a sensitive marker of treatment resistance and of the immune/inflammatory response in that illness. 42, 349–358.

Maes, M. J. J. O. P. M. 2022b. Precision nomothetic medicine in depression research: a new depression model, and new endophenotype classes and pathway phenotypes, and a digital self. 12, 403.

Malhotra, R., Persic, V., Zhang, W., Brown, J., Tao, X., Rosales, L., Thijssen, S., Finkelstein, F. O., Unruh, M. L. & Ikizler, A. 2017. Tryptophan and kynurenine levels and its association with sleep, nonphysical fatigue, and depression in chronic hemodialysis patients. J Renal Nutr, 27, 260–266.

Molfino, A., Imbimbo, G., Amabile, M. I., Ammann, T., Lionetto, L., Salerno, G., Simmaco, M., Chiappini, M. G. & Muscaritoli, M. 2023. Fatigue in Patients on Chronic Hemodialysis: The Role of Indoleamine 2, 3-Dioxygenase (IDO) Activity, Interleukin-6, and Muscularity. Nutrients, 15, 876.

Morris, G. & Maes, M. J. M. B. D. 2013. A neuro-immune model of myalgic encephalomyelitis/chronic fatigue syndrome. 28, 523–540.

Nedergaard, M., Ransom, B. & Goldman, S. A. J. T. I. N. 2003. New roles for astrocytes: redefining the functional architecture of the brain. 26, 523–530.

Ni, M., You, Y., Chen, J. & Zhang, L. J. P. R. 2018. Copper in depressive disorder: A systematic review and meta-analysis of observational studies. 267, 506–515.

Oikawa, H., Hayashi, K. I., Maesawa, C., Masuda, T. & Sobue, K. J. E. C. R. 2010. Expression profiles of nestin in vascular smooth muscle cells in vivo and in vitro. Exp Cell Res, 316, 940–950.

Oweis, A. O., Al-Qarqaz, F., Bodoor, K., Heis, L., Alfaqih, M. A., Almomani, R., Obeidat, M. A. & Alshelleh, S. A. 2021. Elevated interleukin 31 serum levels in hemodialysis patients are associated with uremic pruritus. Cytokine, 138, 155369.

Park, B. S., Lee, H. W., Lee, Y. J., Park, S., Kim, Y. W., Kim, S. E., Kim, I. H., Park, J. H. & Park, K. M. 2020. Serum S100B represents a biomarker for cognitive impairment in patients with end-stage renal disease. Clinical Neurology and Neurosurgery, 195, 105902.

Qawaqzeh, D. T. A., Masa’deh, R., Hamaideh, S. H., Alkhawaldeh, A. & Albashtawy, M. 2023. Factors affecting the levels of anxiety and depression among patients with end-stage renal disease undergoing hemodialysis. Int Urol Nephrol, 1–10.

Sakairi, T., Hiromura, K., Yamashita, S., Takeuchi, S., Tomioka, M., Ideura, H., Maeshima, A., Kaneko, Y., Kuroiwa, T., Nangaku, M., Takeuchi, T. & Nojima, Y. 2007. Nestin expression in the kidney with an obstructed ureter. Kidney International, 72, 307–318.

Sangeetha lakshmi, B., Harini devi, N., Suchitra, M. M., Srinivasa rao, P. & Siva kumar, V. 2018. Changes in the inflammatory and oxidative stress markers during a single hemodialysis session in patients with chronic kidney disease. Ren Fail, 40, 534–540.

Sasaki, Y., Ohsawa, K., Kanazawa, H., Kohsaka, S. & Imai, Y. 2001. Iba1 is an actin-cross-linking protein in macrophages/microglia. Biochem Biophys Res Comm, 286, 292–297.

Savitha, R., Thomas, L., Shetty, M. S. & Kiran, K. J. I. J. O. R. I. P. S. 2020. Comparison of Health Outcomes in Diabetic and Non-Diabetic Patients Undergoing Maintenance Hemodialysis. 11, 2932–2941.

Schouten, R. W., Haverkamp, G. L., Loosman, W. L., Chandie shaw, P. K., Van ittersum, F. J., Smets, Y. F. C., Vleming, L.-J., Dekker, F. W., Honig, A. & Siegert, C. E. H. 2019. Anxiety Symptoms, Mortality, and Hospitalization in Patients Receiving Maintenance Dialysis: A Cohort Study. American Journal of Kidney Diseases, 74, 158–166.

Semaan, V., Noureddine, S. & Farhood, L. 2018. Prevalence of depression and anxiety in end-stage renal disease: A survey of patients undergoing hemodialysis. Appl Nurs Res, 43, 80–85.

Sinuani, I., Beberashvili, I., Averbukh, Z. & Sandbank, J. 2013. Role of IL-10 in the progression of kidney disease. World J Transplant, 3, 91–8.

Song, Y. R., Kim, J. K., Lee, H. S., Kim, S. G. & Choi, E. K. 2020. Serum levels of protein carbonyl, a marker of oxidative stress, are associated with overhydration, sarcopenia and mortality in hemodialysis patients. BMC Nephrol, 21, 281.

Srpova, B., Uher, T., Hrnciarova, T., Barro, C., Andelova, M., Michalak, Z., Vaneckova, M., Krasensky, J., Noskova, L. & Havrdova, E. K. 2021. Serum neurofilament light chain reflects inflammation-driven neurodegeneration and predicts delayed brain volume loss in early stage of multiple sclerosis. Mult Scler J, 27, 52–60.

Steinacker, P., Al shweiki, M. R., Oeckl, P., Graf, H., Ludolph, A. C., Schönfeldt-lecuona, C. & Otto, M. 2021. Glial fibrillary acidic protein as blood biomarker for differential diagnosis and severity of major depressive disorder. J Psychiat Res, 144, 54–58.

Su, H., Lei, C.-T. & Zhang, C. J. F. I. I. 2017. Interleukin-6 signaling pathway and its role in kidney disease: an update. 8, 405.

Su, W., Fang, C., Yang, H.-C., Gu, Y. & Hao, C.-M. J. Z. B. L. X. Z. Z. C. J. O. P. 2008. Expression of nestin in human kidney and its clinical significance. Chinese J Pathol, 37, 309–312.

Timofte, D., Tanasescu, M. D., Balcangiu-Stroescu, A. E., Balan, D. G., Tulin, A., Stiru, O., Vacaroiu, I. A., Mihai, A., Constantin, P. C., Cosconel, C. I., Enyedi, M., Miricescu, D. & Ionescu, D. 2021. Dyselectrolytemia-management and implications in hemodialysis (Review). Exp Ther Med, 21, 102.

Tomioka, M., Hiromura, K., Sakairi, T., Takeuchi, S., Maeshima, A., Kaneko, Y., Kuroiwa, T., Takeuchi, T. & Nojima, Y. 2010. Nestin is a novel marker for renal tubulointerstitial injury in immunoglobulin A nephropathy. Nephrology (Carlton), 15, 568–74.

Vyver, M. V., Beelen, R., De keyser, J., Nagels, G., Van binst, A.-M., Verborgh, C. & D’haeseleer, M. 2018. Plasma citrulline levels are increased in patients with multiple sclerosis. J Neurolog Sci, 387, 174–178.

Wang, D. D. & Bordey, A. J. P. I. N. 2008. The astrocyte odyssey. 86, 342–367.

Wang, P., Qin, D. & Wang, Y.-F. J. F. I. M. N. 2017a. Oxytocin rapidly changes astrocytic GFAP plasticity by differentially modulating the expressions of pERK 1/2 and protein kinase A. 10, 262.

Wang, Q., Jie, W., Liu, J.H., Yang, J. M. & Gao, T. M. 2017b. An astroglial basis of major depressive disorder? An overview. Glia, 65, 1227–1250.

WĄsik, N., SokóŁ, B., HoŁysz, M., MaŃko, W., Juszkat, R., JagodziŃski, P. P. & Jankowski, R. J. A. N. 2020. Serum myelin basic protein as a marker of brain injury in aneurysmal subarachnoid haemorrhage. 162, 545–552.

Watanabe, M., Nakamura, Y., Michalak, Z., Isobe, N., Barro, C., Leppert, D., Matsushita, T., Hayashi, F., Yamasaki, R. & Kuhle, J. J. N. 2019. Serum GFAP and neurofilament light as biomarkers of disease activity and disability in NMOSD. Neurol, 93, e1299–e1311.

Yang, Z. & Wang, K. K. J. T. I. N. 2015. Glial fibrillary acidic protein: from intermediate filament assembly and gliosis to neurobiomarker. 38, 364–374.

Yoong, R. K., Mooppil, N., Khoo, E. Y., Newman, S. P., Lee, V. Y., Kang, A. W. & Griva, K. 2017. Prevalence and determinants of anxiety and depression in end stage renal disease (ESRD). A comparison between ESRD patients with and without coexisting diabetes mellitus. J Psychosom Res, 94, 68–72.

Zachrisson, O., Regland, B., Jahreskog, M., Kron, M. & Gottfries, C. G. 2002. A rating scale for fibromyalgia and chronic fatigue syndrome (the FibroFatigue scale). J Psychosom Res, 52, 501–509.

Zhang, J., Sun, X., Zheng, S., Liu, X., Jin, J., Ren, Y. & Luo, J. 2014. Myelin basic protein induces neuron-specific toxicity by directly damaging the neuronal plasma membrane. PLoS One, 9, e108646.

